# The UK Divide: Does having a Pembrolizumab-Chemotherapy option in head and neck cancer matter? Real-world experience of first-line palliative pembrolizumab monotherapy and pembrolizumab-chemotherapy combination in Scotland

**DOI:** 10.1101/2023.06.08.23290541

**Authors:** Alekh Thapa, Anna Cowell, Adam Peters, David J Noble, Allan James, Carolynn Lamb, Derek Grose, Saurabh Vohra, Stefano Schipani, Karen Mactier, Joanna Mackenzie, Devraj Srinivasan, Kirsten Laws, Rafael Moleron, Paddy Niblock, Feng Yi Soh, Claire Paterson, Christina Wilson

## Abstract

**Objectives:** The Scottish Medical Consortium recently approved first-line pembrolizumab monotherapy or in combination with chemotherapy for head and neck squamous cell carcinoma (HNSCC) in the palliative setting, contrasting with the decision made by the National Institute for Health and Care Excellence who approved monotherapy alone in England and Wales. We aimed to provide real-world performance data for first-line pembrolizumab-containing treatments for head and neck squamous cell carcinoma (HNSCC) in the palliative setting in Scotland.

**Materials and Methods:** We analysed the electronic records of patients who initiated pembrolizumab-containing treatment between 01/03/2020–30/09/2021. Outcomes included overall survival (OS), progression-free survival (PFS), duration of response (DOR), disease control rate (DCR). Data were compared with the KEYNOTE-048 study and clinical factors were evaluated for association with survival.

**Results:** Our cohort included 91 patients (median follow-up 10.8 months). Patient characteristics were similar to the KEYNOTE-048 study though our cohort had a higher proportion of patients with newly diagnosed, non-metastatic disease.

For patients receiving monotherapy (n=76), 12-month and 24-month OS was 45% and 27%, respectively. For patients receiving pembrolizumab-chemotherapy (n=15), 12-month OS was 60% (24-month OS had not yet been reached). Experiencing ≥1 irAE (versus no irAEs), of any grade, was associated with favourable OS and PFS for patients receiving monotherapy in both univariable log-rank analysis (median OS 17.4 months versus 8.6 months, respectively, P=0.0033; median PFS 10.9 months versus 3.0 months, respectively, P<0.0001) and multivariable analysis (Cox proportional hazards regression: OS HR: 0.31, P=0.0009; PFS HR: 0.17, P<0.0001).

**Conclusion:** Our real-world data support the KEYNOTE-048 study findings and the value of combination treatment options. Additionally, our data show irAEs of any grade, as reported in routine clinical records, are associated with better outcomes in this patient group, adding to the growing body of evidence showing irAEs are generally a positive marker of PD-L1 inhibitor response.

## Introduction

Palliative treatments are central to the management of head and neck squamous cell cancers (HNSCC) due to the predominance of advanced disease at diagnosis and the high levels of recurrence and comorbidities [1, 2]. Within Scotland, only 56% of all patients diagnosed survive beyond 5 years after diagnosis, with this survival rate remaining relatively unchanged for the past two decades [3].

The advent of immune checkpoint inhibitors has represented a paradigm shift in cancer care and offers hope for improving these stubbornly static survival outcomes. In particular, the programmed death-1 (PD-1)-mediated inhibition of CD8+ T cell anti-tumour activity has been identified as a critical axis for tumour immune surveillance escape [4]. The recent KEYNOTE-048 Phase 3 randomised controlled trial (RCT) demonstrated the efficacy of pembrolizumab as a first-line treatment for recurrent and metastatic (R/M) HNSCC with international guidelines now recommending pembrolizumab, with or without chemotherapy, as the standard of care first-line treatment for R/M HNSCC with a PD-L1 combined positive score (CPS) ≥1 [2, 5–7].

In Scotland, pembrolizumab received short-term approval for use without PD-L1 CPS testing as an alternative to immunosuppressing cytotoxic chemotherapy during the COVID-19 pandemic [8]. Since September 2020, it has had full Scottish Medical Consortium approval, with or without chemotherapy, in the first-line palliative setting, for HNSCC with PD-L1 CPS ≥1 [9]. This is at odds with National Institute for Health and Care Excellence (NICE) guidance in England and Wales which recommends pembrolizumab monotherapy only in the same patient cohort [10]. Clinicians in NHS England have already voiced concerns about this decision [11]. Notably, in the KEYNOTE-048 study the overall survival (OS) curve for patients receiving pembrolizumab monotherapy was below the active control in the first 7–8 months of treatment before crossing over, with objective response rate (ORR) also lower in the pembrolizumab monotherapy arm [5]. In contrast, the OS curve for patients receiving pembrolizumab-chemotherapy was similar to active control in the same time period, and ORR was similar to active control. Furthermore, post-hoc analysis of the KEYNOTE-048 study has suggested that patients with moderate levels of PD-L1 may benefit the most from a pembrolizumab-chemotherapy combination rather than pembrolizumab monotherapy [12].

Whilst strict inclusion and exclusion criteria used within RCTs allow for minimally biased evidence, the study population differs from real-world patient populations, tending to be younger and fitter [13]. Real-world studies are valuable in assessing the efficacy and toxicity of new treatments in routine clinical practice and also allow analysis of factors influencing treatment response [14, 15]. For example, immune-related adverse events (irAEs) are predictive of better outcomes for patients receiving PD-L1 inhibitors in several cancers such as the second-line palliative setting with Nivolumab in HNSCC [16–19]. Such real-world studies are lacking for first-line palliative pembrolizumab-containing regimens for HNSCC.

To address this gap in the literature, we conducted a national, multi-centre, retrospective cohort study with two aims: to provide real-world performance data and to explore factors predicting clinical outcomes using univariable and multivariable analyses.

## Patients and methods

### Study Design and Participants

We conducted a retrospective cohort study using electronic records for patients treated at five tertiary cancer care centres covering the entire Scottish population of 5.3 million people. The study collected anonymised health service clinical practice data and was approved by NHS Information Governance. REporting of studies Conducted using Observational Routinely collected health Data (RECORD) guidelines were followed [20].

Patients were identified by searching the local Chemotherapy Electronic Prescribing and Administration Systems (CEPAS). Linked records were identified in the local radiotherapy records, electronic clinical records and national radiology records using unique National Health Service Scotland patient identifier numbers (Figure 1). A standardised data collection form was used for all centres and anonymised data was collated at a single site for analysis.

**Figure 1.**
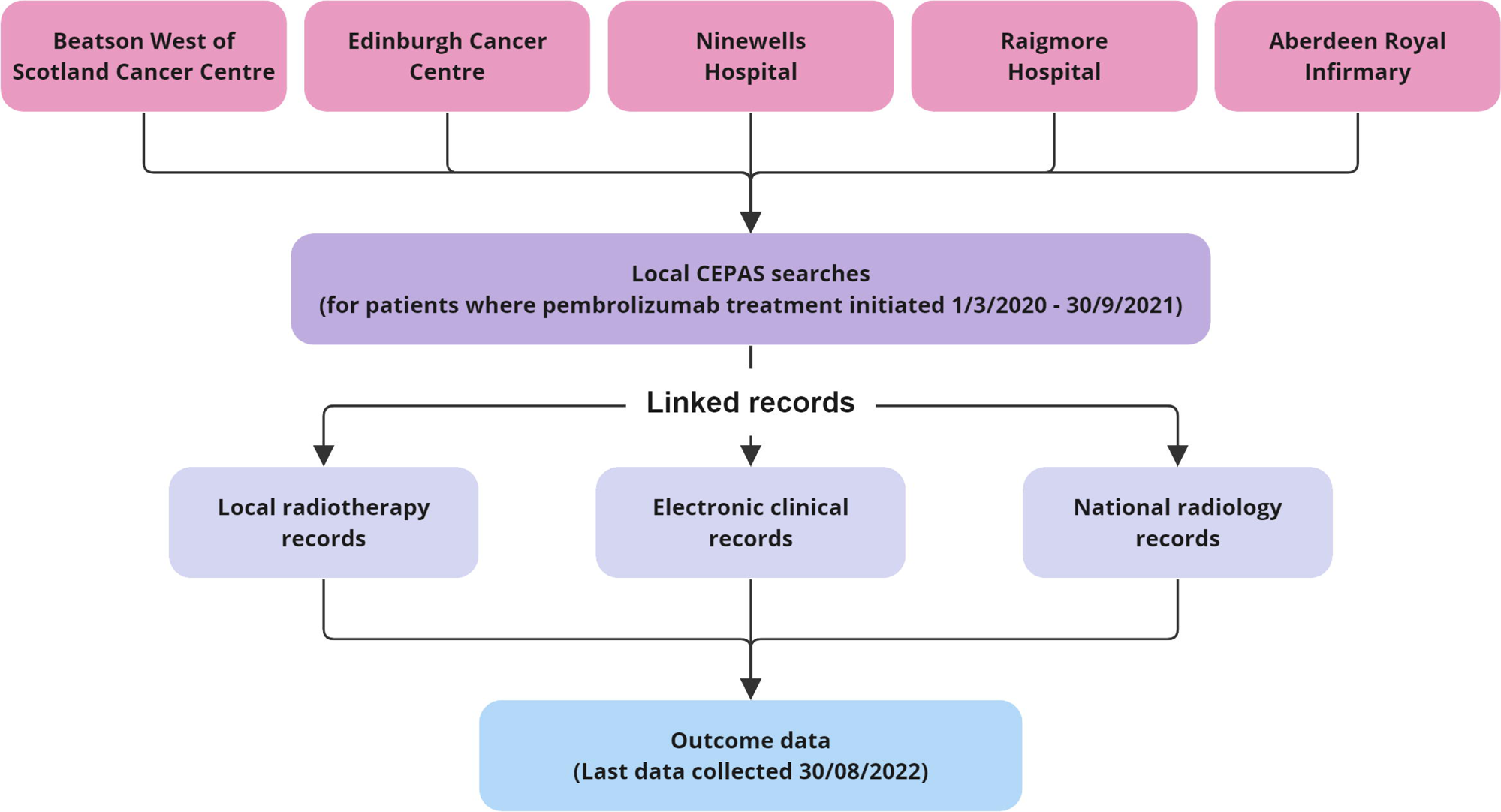
Record linkage flow diagram. Cancer centres involved in the study are shown at the top. CEPAS, Chemotherapy Electronic Prescribing and Administration System.

Eligible patients had initiated pembrolizumab-containing treatment between 1^st^ March 2020 to 30^th^ September 2021 as first-line palliative therapy for HNSCC. Patients received either pembrolizumab monotherapy (200 mg 3-weekly or 400 mg 6-weekly intravenously [IV]) or pembrolizumab-chemotherapy (pembrolizumab 200 mg IV Day 1, cisplatin 75–100mg/m² or carboplatin AUC 5mg/mL/min IV Day 1, and 5-Fluorouracil 750–1000mg/m² IV Day 1–4) every 21 days for up to 6 cycles followed by pembrolizumab maintenance monotherapy as described. Patients continued treatment for a maximum of two years, until disease progression or unacceptable toxicity. Patients were excluded from this analysis if they received pembrolizumab as part of a clinical study.

Patient characteristics collected were age, gender, Eastern Cooperative Oncology Group (ECOG) performance status (PS), smoking history and any relevant comorbidities. Disease characteristics collected were primary tumour subsite, pathology (including p16 status for oropharyngeal and cancers of unknown primary subsites, and PD-L1 CPS), and treatment history. Follow-up data collected included irAEs, additional treatments received during and after pembrolizumab, radiological response, and the date and cause of death. Adverse reactions were graded according to Common Terminology Criteria for Adverse Events (CTCAE) v5.

### Statistical Analysis

OS, PFS and duration of response (DOR) were estimated using Kaplan-Meier analysis. OS was defined as the time from cycle 1 of pembrolizumab to the date of death from any cause. PFS was defined as the time from cycle 1 of pembrolizumab to the date of radiological progression, decline in performance-related treatment cessation, or death from any cause, whichever came first. Patients were censored for all survival analyses if still alive at the end of the follow-up period or if lost to follow-up. DOR was defined, as per KEYNOTE-048 [5], as the length of time from radiological response to progression, decline in performance requiring treatment cessation or death for patients with radiological response at any time whilst receiving pembrolizumab-containing treatment. Log-rank test was used for survival curve comparative analysis.

The responsible treating clinician defined radiological response, using both clinical and radiological evaluations. Where the treating clinical teams stated the disease as responding to treatment, we categorised this as “response”. Where it was stated there was a mixed response or only some of the disease locations were responding, we categorised this as “mixed response”. Where the treating clinical teams stated the disease was stable or there was no change from the last cross-sectional imaging reports, we categorised this as “stable disease”. Where it was stated that the disease was progressing or no follow-up cross-sectional imaging was performed due to deterioration PS or death, we categorised this as “progressive disease”. Best radiological response during treatment with pembrolizumab was reported.

For disease control rate (DCR) analysis, DCR was defined as the proportion of patients with a best radiological response categorised as response, mixed response or stable disease. Fisher’s exact test was used to analyse differences in DCR.

Cox proportional hazards regression was used for univariable and multivariable analyses. For the multivariable analysis, confounding factors were selected for inclusion if associated with differential overall survival in KEYNOTE-048 and potentially with irAEs [5, 21]. All statistical analysis was performed using GraphPad PRISM v9.5.1.

## Results

### Patient characteristics

Of our 91-patient cohort, two (2.2%) patients were lost to follow-up. Median follow-up was 10.8 months (range 0.6–25.3 months), 10.7 months (range 1.0–25.3 months) and 11.7 months (range 0.6–18.6 months) for the whole cohort, patients treated with pembrolizumab monotherapy, and patients treated with pembrolizumab-chemotherapy, respectively.

Baseline characteristics are shown in Table 1. PD-L1 CPS was not measured in 21 (27.6%) patients receiving pembrolizumab monotherapy during the short-term COVID-19 pandemic-related approval period when this was not required. Two patients with nasopharyngeal tumours were included, both were EBV-negative/p16-positive HNSCC. Whilst patients with EBV-driven nasopharyngeal cancer represent a distinct disease entity, the patients included in our cohort were felt to represent a mucosal SCC, in keeping with those found at other sub-sites.

**Table 1.** Baseline demographics in the present study (Scottish study) and the KEYNOTE-048 study [5]. In the present study, a p16 result was only recorded for oropharyngeal and unknown primary site tumours. CPS, combined positive score; PD-L1, programmed death-ligand 1.

|  | Scottish study |  | KEYNOTE-048 |  |
| --- | --- | --- | --- | --- |
|  | Pembrolizumab monotherapy | Pembrolizumab -chemotherapy | Pembrolizumab monotherapy | Pembrolizumab -chemotherapy |
|  | n = 76 | n = 15 | n = 301 | n = 281 |
| Age, median years (range) |  |  |  |  |
|  | 66 (38–86) | 53 (34–76) | 62 (56–68) | 61 (55–68) |
| Age, years, n (%) |  |  |  |  |
| ≤50 | 5 (6.6) | 4 (26.7) |  |  |
| 50–70 | 51 (67.1) | 9 (60.0) |  |  |
| >70 | 20 (26.3) | 2 (13.3) |  |  |
| Gender, n (%) |  |  |  |  |
| Male | 55 (72.4) | 8 (53.3) | 250 (83)* | 224 (80)* |
| Female | 21 (27.6) | 7 (46.7) | 51 (17)* | 57 (20)* |
| ECOG performance status, n (%) |  |  |  |  |
| 0 | 15 (19.7) | 6 (40.0) | 118 (39) | 110 (39) |
| 1 | 56 (73.7) | 9 (60.0) | 183 (61) | 171 (61) |
| 2 | 5 (6.6) | 0 (0.0) |  |  |
| Smoking status, n (%) |  |  |  |  |
| Current or former | 67 (88.2) | 12 (80.0) | 239 (79) | 224 (80) |
| Never | 9 (11.8) | 3 (20.0) | 62 (21) | 57 (20) |
| p16 status, n (%) |  |  |  |  |
| Positive | 11 (14.5) | 4 (26.7) | 63 (21) | 60 (21) |
| Negative | 17 (22.4) | 2 (13.3) |  |  |
| Not done/Not applicable | 48 (63.2) | 9 (60.0) |  |  |
| PD-L1 CPS, n (%) |  |  |  |  |
| ≥1 – <20 | 38 (50.0) | 8 (53.3) | 124 (41) | 116 (41) |
| ≥20 | 17 (22.4) | 7 (46.7) | 133 (44) | 126 (45) |
| Not measured | 21 (27.6) | 0 (0.0) |  |  |
| Disease status, n (%) |  |  |  |  |
| Newly diagnosed, non-metastatic | 27 (35.5) | 6 (40.0) | 3 (1) | 4 (1) |
| Locoregional recurrence<sup>†</sup> | 24 (31.6) | 7 (46.7) | 82 (27) | 76 (27) |
| Metastatic | 25 (32.9) | 2 (13.3) | 216 (72) | 201 (72) |
| Primary tumour site, n (%) |  |  |  |  |
| Oropharynx | 26 (34.2) | 6 (40.0) | 113 (38) | 113 (40) |
| Oral cavity | 12 (15.8) | 7 (46.7) | 82 (27) | 82 (29) |
| Hypopharynx | 20 (26.3) | 0 (0.0) | 38 (13) | 44 (16) |
| Larynx | 11 (14.5) | 0 (0.0) | 74 (25) | 46 (16) |
| Nasopharynx | 1 (1.3) | 1 (6.7) |  |  |
| Sinus | 3 (3.9) | 1 (6.7) |  |  |
| Unknown primary site | 3 (3.9) | 0 (0.0) |  |  |
| Platinum therapy received, n (%) |  |  |  |  |
| Cisplatin | N/A | 14 (93.3) |  | 121 (43) |
| Carboplatin | N/A | 1 (6.7) |  | 160 (57) |

*\*KEYNOTE-048 study reported the sex of participants. <sup>†</sup>Recurrent disease at primary site and/or regional lymph nodes.*

### irAE toxicity

Acknowledging the limitations of retrospective toxicity analyses, we focused on grade ≥3 irAE toxicities as shown in Table 2, as felt those were most likely to be recorded robustly in routine clinical records. For those receiving pembrolizumab monotherapy, six patients (7.9%) experienced an irAE graded ≥3 (total number toxicities=7). The most frequent toxicities were hepatitis (5.3%) and hypoadrenalism (2.6%).

**Table 2.** Immune-related adverse event (grade ≥3) profile from our study and the KEYNOTE-048 trial. GI, gastrointestinal; irAE, immune-related adverse event.

|  | Scottish study |  | KEYNOTE-048* |  |
| --- | --- | --- | --- | --- |
|  | Pembrolizumab monotherapy | Pembrolizumab -chemotherapy | Pembrolizumab monotherapy | Pembrolizumab -chemotherapy |
| Grade $\geq 3$ irAE, n (%) | n = 76 | n = 15 | n = 300 | n = 276 |
| GI disorders | 4 (5.3) | 1 (6.7) |  |  |
| Hepatitis | 3 (3.9) | 0 (0.0) | 2 (0.7) | 1 (0.4) |
| Diarrhoea | 1 (1.3) | 1 (6.7) | 2 (0.7) | 8 (2.9) |
| Pancreatitis | 0 (0.0) | 0 (0.0) | 0 (0.0) | 1 (0.4) |
| Endocrine disorders | 2 (2.6) | 0 (0.0) |  |  |
| Hypoadrenalism | 2 (2.6) | 0 (0.0) | 1 (0.3) | 0 (0.0) |
| Hypophysitis | 0 (0.0) | 0 (0.0) | 1 (0.3) | 1 (0.4) |
| Hyperthyroidism | 0 (0.0) | 0 (0.0) | 1 (0.3) | 0 (0.0) |
| Systemic, n (%) | 0 (0.0) | 0 (0.0) |  |  |
| Fatigue | 0 (0.0) | 0 (0.0) | 9 (3.0) | 20 (7.2) |
| Dermatological disorders | 1 (1.3) | 1 (6.7) |  |  |
| Rash | 1 (1.3) | 1 (6.7) | 2 (0.7) | 1 (0.4) |
| Cardiovascular disorders | 0 (0.0) | 0 (0.0) |  |  |
| Myocarditis | 0 (0.0) | 0 (0.0) | 0 (0.0) | 1 (0.4) |
| Respiratory disorders | 0 (0.0) | 0 (0.0) |  |  |
| Pneumonitis | 0 (0.0) | 0 (0.0) | 5 (1.7) | 5 (1.8) |
| Renal disorders | 0 (0.0) | 0 (0.0) |  |  |
| Nephritis | 0 (0.0) | 0 (0.0) | 2 (0.7) | 0 (0.0) |
| Neurological disorders | 0 (0.0) | 0 (0.0) |  |  |
| Encephalitis | 0 (0.0) | 0 (0.0) | 1 (0.3) | 0 (0.0) |

*\*KEYNOTE-048 reported selected adverse events based on a frequency threshold or if they were of interest to investigators. Total numbers within each category have thus been omitted as this is not comparable to our data.*

For pembrolizumab-chemotherapy, two patients (13.3%) experienced an irAE graded ≥3 (total number toxicities=2). These were diarrhoea and rash.

### Efficacy

For patients receiving pembrolizumab monotherapy, 12-month and 24-month OS was 45% and 27%, respectively (Figure 1a), and median OS was 11.4 months. The 12-month and 24-month PFS were 22% and 14%, respectively (Figure 1c), and median PFS was 6.4 months. Median DOR was 13.3 months (Figure 1e) with a DCR of 56.6% (35.5% response, 11.8% mixed response and 9.2% stable disease) (Figure 1g).

For patients receiving pembrolizumab-chemotherapy, 12-month OS was 60% (Figure 1b). PFS was 20% at 12 months and median PFS was 4.5 months (Figure 1d). Median DOR was 7.3 months with a DCR of 60.0% (53.3% response, 6.7% mixed response and 0.0% stable disease, Figure 1h). Median OS, 24-month OS and PFS had not yet been reached for these patients.

### Association of irAEs and outcome

In line with previous real-world studies, we included all-grade irAEs when analysing for associations with differential OS and PFS [16, 17, 22–24]. The profile of all-grade irAEs in our cohort are shown in Table S1. Kaplan-Meier and Log-rank analysis showed significantly better OS (median OS 17.4 versus 8.6 months, respectively, P=0.0033) and PFS (median PFS 10.9 versus 3.0 months, respectively, P<0.0001) for patients who experienced ≥1 irAE and received pembrolizumab monotherapy compared with experiencing none (Figure 2a and 2c). Similar but non-significant trends were observed for the pembrolizumab-chemotherapy group (Figure 2).

**Figure 2.**
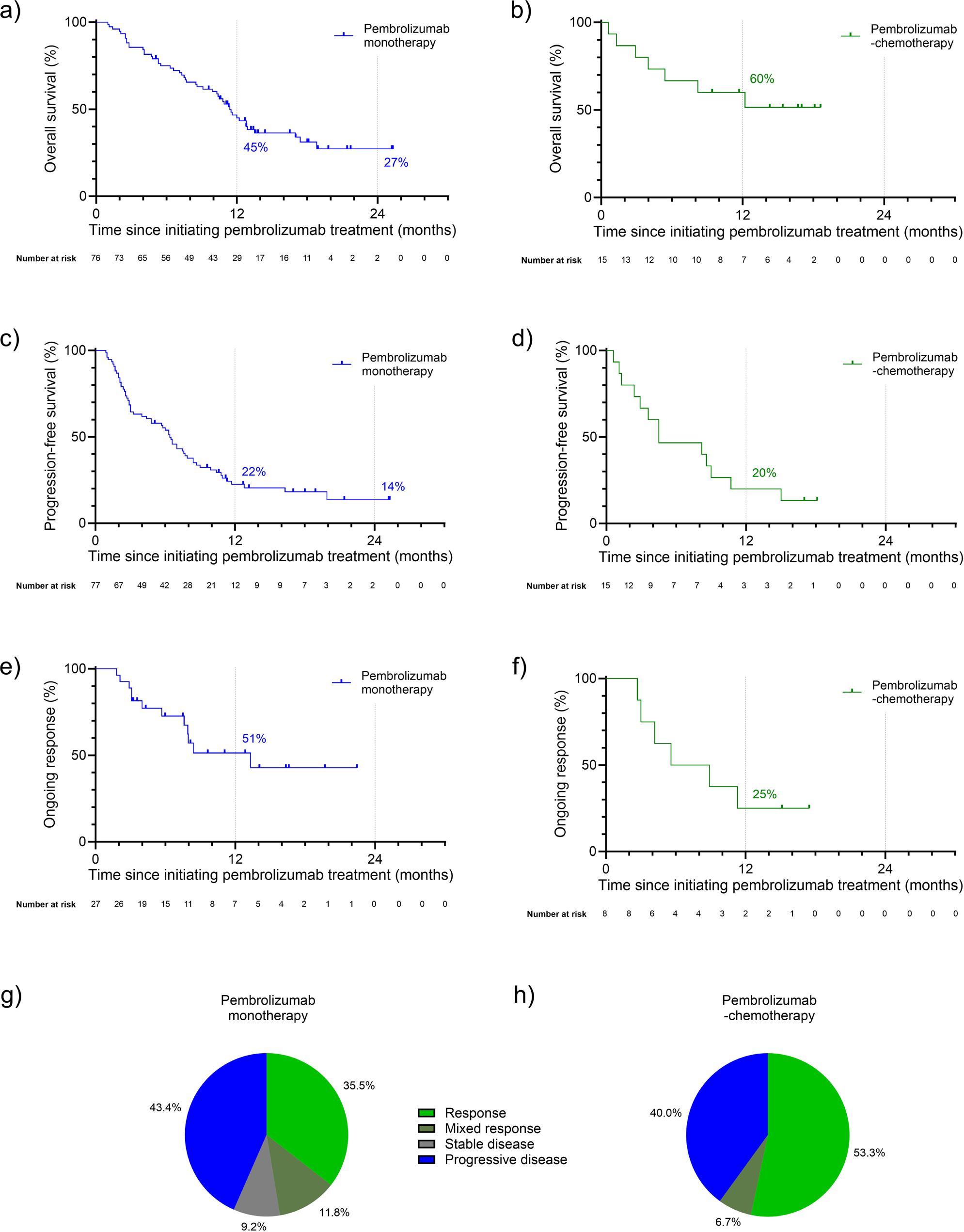
Kaplan-Meier survival curves for overall survival, progression-free and duration of response in patients receiving pembrolizumab monotherapy (a), c) and e), respectively) or with chemotherapy (b), d) and f), respectively). Where reached, percentage survival at 12- and 24-month are shown. Best radiological response during treatment for patients receiving g) pembrolizumab monotherapy or h) with chemotherapy. No patients receiving pembrolizumab-chemotherapy had the best radiological response categorised as stable disease.

Best radiological response stratified by experiencing ≥1 irAE is shown in Figures 2e and 2f. For patients experiencing ≥1 irAE of any grade and receiving pembrolizumab monotherapy, DCR was significantly greater (P<0.0001) at 89.3% (64.3% response, 17.9% mixed response and 7.1% stable disease) versus 37.5% for no irAEs (18.8% response, 8.3% mixed response and 10.4% stable disease).

Similarly, for patients experiencing ≥1 irAE of any grade and receiving pembrolizumab-chemotherapy, DCR was significantly greater (P=0.0278) at 100.0% (83.3% response, 16.7% mixed response and 0.0% stable disease) versus 33.3% for no irAEs (33.3% response, 0.0% mixed response and 0.0% stable disease).

**Figure 3.**
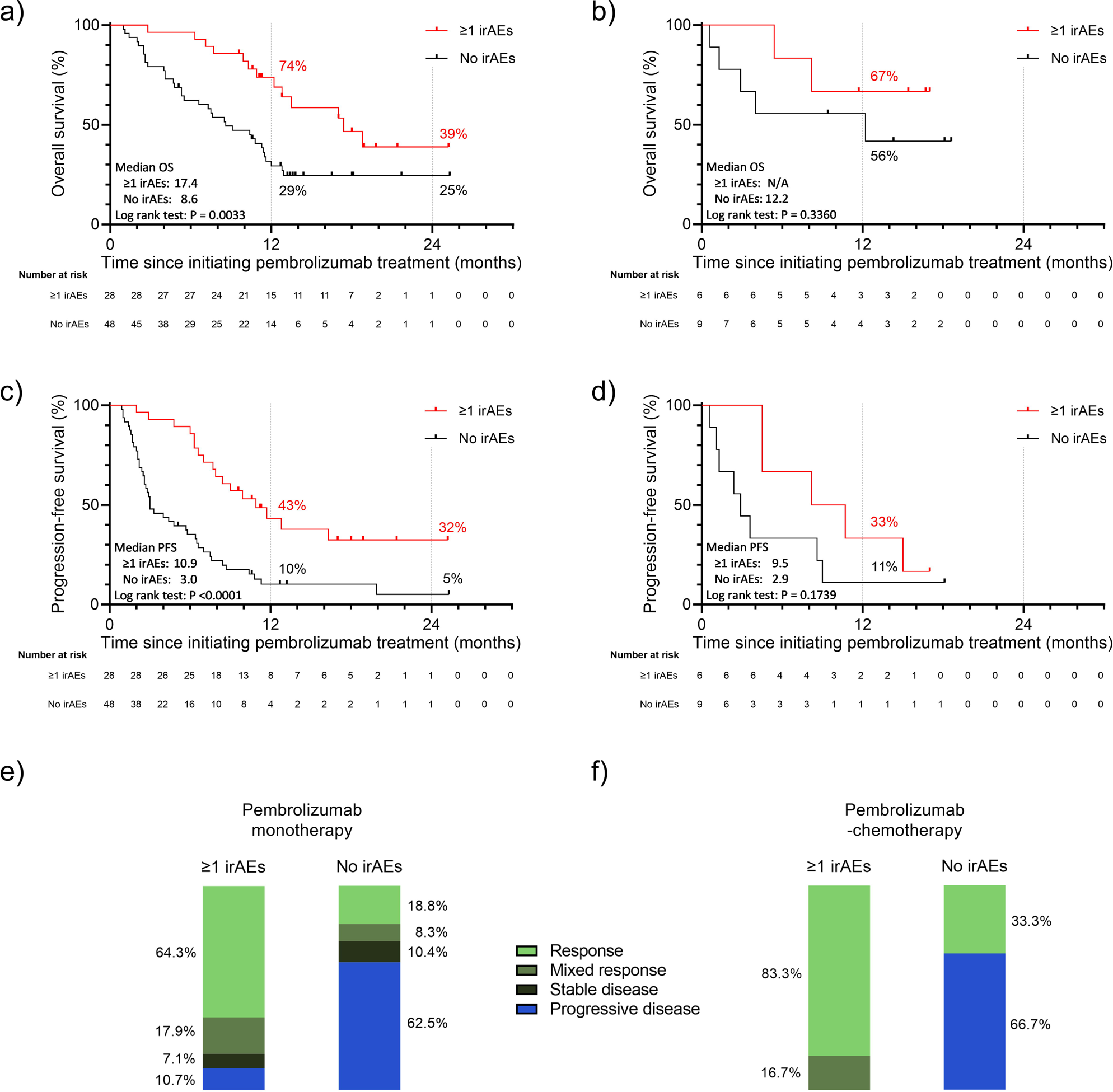
Kaplan-Meier survival curves for a) overall survival and c) progression-free survival in patients receiving pembrolizumab monotherapy, and b) overall survival and d) progression-free survival in patients receiving pembrolizumab with chemotherapy, stratified by the presence or absence of irAEs of any grade. Best radiological response during treatment as judged by the treating clinical team for patients receiving e) pembrolizumab monotherapy and f) pembrolizumab-chemotherapy, who experienced 1 or more irAE of any grade (left bars) or no irAE (right bars). No patients receiving pembrolizumab-chemotherapy had the best radiological response categorised as stable disease. irAEs, immune-related adverse events; N/A, not applicable; OS, overall survival; PFS, progression-free survival.

### Identifying other clinical factors associated with OS and PFS

To investigate other clinical factors for association with differential OS or PFS in patients receiving pembrolizumab monotherapy, we performed univariable Cox proportional hazards regression (Table 3).

**Table 3.** Univariable analysis of factors potentially associated with OS and PFS for patients receiving pembrolizumab monotherapy. Multivariable analysis adjusting for factors potentially confounding the association between experiencing ≥1 irAE and OS ad PFS. Statistically significant P values are shown in bold and underlined. The most frequent category was chosen as the reference. CI, confidence interval; CPS, combined positive score; Dx, diagnosis; HR, hazard ratio; irAEs, immune-related adverse events; OS, overall survival; PD-L1, programmed death-ligand 1; PFS, progression-free survival; PS, performance score; XRT, radiotherapy.

| Factor | OS |  |  |  |  |  | PFS |  |  |  |  |  |
| --- | --- | --- | --- | --- | --- | --- | --- | --- | --- | --- | --- | --- |
|  | Univariable analysis |  |  | Multivariable analysis |  |  | Univariable analysis |  |  | Multivariable analysis |  |  |
|  | HR | (95% CI) | P value | HR | (95% CI) | P value | HR | (95% CI) | P value | HR | (95% CI) | P value |
| $\geq 1$ irAE any grade (reference: No) | | | | | | | | | | | | |
| Yes | 0.39 | (0.20–0.73) | **<u>0.0045</u>** | 0.31 | (0.15–0.60) | **<u>0.0009</u>** | 0.32 | (0.18–0.56) | **<u><0.0001</u>** | 0.17 | (0.08–0.33) | **<u><0.0001</u>** |
| $\geq 1$ irAE grade $\geq 3$ (reference: No) | | | | | | | | | | | | |
| Yes | 0.72 | (0.21–1.78) | 0.5258 |  |  |  | 0.49 | (0.15–1.12) | 0.1745 |  |  |  |
| Age (reference: $\geq 65$ years) | | | | | | | | | | | | |
| <65 years | 0.91 | (0.48–1.64) | 0.7515 | 1.36 | (0.53–3.22) | 0.5038 | 0.89 | (0.51–1.50) | 0.6610 | 1.16 | (0.50–2.63) | 0.7190 |
| Gender (reference: Male) |  |  |  |  |  |  |  |  |  |  |  |  |
| Female | 1.61 | (0.88–2.85) | 0.1131 |  |  |  | 1.62 | (0.93–2.76) | 0.0791 |  |  |  |
| ECOG PS at cycle 1 (reference: 1) |  |  |  |  |  |  |  |  |  |  |  |  |
| 0 | 0.55 | (0.23–1.17) | 0.1515 | 0.46 | (0.17–1.10) | 0.1002 | 0.62 | (0.30–1.19) | 0.1791 | 0.50 | (0.20–1.13) | 0.1169 |
| 2 | 3.38 | (1.13–8.22) | **<u>0.0141</u>** | 3.95 | (1.06–13.47) | **<u>0.0311</u>** | 1.89 | (0.65–4.37) | 0.1801 | 1.91 | (0.52–6.31) | 0.3026 |
| Smoking status (reference: Current or former) |  |  |  |  |  |  |  |  |  |  |  |  |
| Never | 1.64 | (0.67–3.46) | 0.2312 |  |  |  | 1.27 | (0.56–2.54) | 0.5265 |  |  |  |
| p16 status (reference: negative) |  |  |  |  |  |  |  |  |  |  |  |  |
| Positive | 1.04 | (0.32–3.03) | 0.9383 | 0.88 | (0.22–3.09) | 0.8401 | 1.04 | (0.39–2.59) | 0.9324 | 0.67 | (0.19–2.14) | 0.5156 |
| Not done/Not applicable | 1.84 | (0.92–4.10) | 0.1037 | 2.31 | (1.03–5.71) | 0.0528 | 1.63 | (0.88–3.24) | 0.1384 | 2.34 | (1.09–5.43) | **<u>0.0367</u>** |
| PD-L1 CPS<br>(reference: $\geq 1$ to $<20$) | | | | | | | | | | | | |
| $\geq 20$ | 1.38 | (0.66–2.72) | 0.3719 | 1.45 | (0.67–3.02) | 0.3247 | 1.65 | (0.86–3.06) | 0.1167 | 1.73 | (0.87–3.36) | 0.1096 |
| Not measured | 0.89 | (0.44–1.74) | 0.7395 | 0.61 | (0.29–1.25) | 0.1889 | 0.80 | (0.42–1.47) | 0.4862 | 0.52 | (0.25–1.02) | 0.0650 |
| Disease status<br>(reference: Newly diagnosed, non-metastatic) |  |  |  |  |  |  |  |  |  |  |  |  |
| Locoregional recurrence* | 0.60 | (0.28–1.24) | 0.1790 | 0.50 | (0.22–1.10) | 0.0925 | 0.76 | (0.40–1.43) | 0.3939 | 0.61 | (0.28–1.26) | 0.1896 |
| Metastatic | 1.06 | (0.55–2.03) | 0.8605 | 1.16 | (0.56–2.34) | 0.6800 | 1.46 | (0.79–2.68) | 0.2252 | 2.29 | (1.11–4.74) | **0.0244** |
| Primary tumour site<br>(reference: Oropharynx) |  |  |  |  |  |  |  |  |  |  |  |  |
| Oral cavity | 1.37 | (0.54–3.26) | 0.4829 |  |  |  | 1.40 | (0.62–2.98) | 0.3934 |  |  |  |
| Hypopharynx | 1.74 | (0.82–3.72) | 0.1468 |  |  |  | 1.56 | (0.81–3.03) | 0.1819 |  |  |  |
| Larynx | 1.68 | (0.59–4.27) | 0.2924 |  |  |  | 1.24 | (0.51–2.76) | 0.6193 |  |  |  |
| Nasopharynx | 19.24 | (0.98–127.30) | **0.0083** |  |  |  | 18.16 | (0.93–118.30) | **0.0093** |  |  |  |
| Sinus | 3.25 | (0.74–10.17) | 0.0671 |  |  |  | 1.58 | (0.37–4.68) | 0.4653 |  |  |  |
| Unknown primary site | 1.37 | (0.21–4.98) | 0.6818 |  |  |  | 0.73 | (0.12–2.52) | 0.6665 |  |  |  |
| Primary XRT<br>(reference: No) |  |  |  |  |  |  |  |  |  |  |  |  |
| Yes | 1.04 | (0.57–1.85) | 0.8966 |  |  |  | 1.07 | (0.63–1.78) | 0.8104 |  |  |  |
| Palliative XRT received before/during pembrolizumab (reference: No)<sup>†</sup> |  |  |  |  |  |  |  |  |  |  |  |  |
| Yes | 1.16 | (0.61–2.11) | 0.6291 |  |  |  | 1.44 | (0.81–2.47) | 0.1960 |  |  |  |
| Time from Dx to Pembrolizumab<br>(reference: $<12$ months) | | | | | | | | | | | | |
| $\geq 12$ months | 0.88 | (0.48–1.57) | 0.6666 | | | | 0.83 | (0.49–1.40) | 0.4961 | | | |

*\*Recurrent disease at primary site and/or regional lymph nodes; <sup>†</sup>Palliative radiotherapy regimes were variable and ranged from a single 8 Gy fraction to 50 Gy in 25 fractions. Additional detail is available in Table S2.*

Our cohort included a substantial proportion of patients with newly-diagnosed, non-metastatic tumours (27 patients [35.5%] of those receiving pembrolizumab monotherapy), a group included but with low representation in the KEYNOTE-048 study (1%) [5]. When comparing OS and PFS for this group versus patients with locoregional recurrence or metastatic disease, using univariable Cox proportional hazards regression, we found similar survival outcomes (Table 3).

### Multivariable survival analysis

Since potential factors have been associated with differential outcomes in the KEYNOTE-048 study subgroup analysis [5, 21], We controlled for these possible confounding covariables using multivariable analysis (Table 3). When accounting for age, ECOG PS, disease status (metastatic; locoregional recurrence; newly-diagnosed, non-metastatic), p16 status and PD-L1 CPS, experiencing ≥1 irAE remained highly significantly associated with both OS (HR: 0.31 [95% confidence interval: 0.15–0.60], P=0.0009) and PFS (HR: 0.17 [95% confidence interval: 0.08–0.33], P<0.0001).

## Discussion

Our national retrospective cohort study reports the real-world performance of pembrolizumab monotherapy and pembrolizumab-chemotherapy as first-line palliative treatment for patients with HNSCC in a Scotland-based population and adds to the limited worldwide real-world data for this treatment.

Within our cohort, we found 12-month and 24-month OS was 45% and 27%, respectively, for patients who had received pembrolizumab monotherapy, with a median OS of 11.4 months. This is reassuringly comparable to the KEYNOTE-048 study where 12-month and 24-month OS (the primary endpoint) was 49% and 27%, respectively, with a median OS of 11.6 months despite our cohort contained fewer PS 0 patients (Table 4) [5]. For patients in our cohort who had received pembrolizumab-chemotherapy, 12-month OS was 60%, with 24-month and median OS yet to be reached. Again, this was broadly similar to the KEYNOTE-048 study where 12-month OS was 53% for patients receiving combination treatment (Table 4). Similarly, 12-month PFS for patients receiving pembrolizumab monotherapy or pembrolizumab-chemotherapy was also comparable to the same endpoints in the KEYNOTE-048 study (Table 4). Three other studies have reported the real-world OS for patients receiving pembrolizumab with or without chemotherapy as first-line palliative treatment for HNSCC (two Japanese and one USA population study). All reported similar results to our cohort [25–27]. One study, of a Japanese cohort, reported 12-month OS was 51.9% and 58.8% for patients receiving pembrolizumab monotherapy and pembrolizumab-chemotherapy, respectively [26]. The second Japanese study did not stratify analyses by treatment type and reported a 12-month OS of 64.5% for all patients receiving any form of pembrolizumab treatment [25]. The third study, of a USA cohort, reported a median OS of 8.8 months and found no significant difference between their observational and reconstructed survival data from the KEYNOTE-048 study [27]. Overall, our data suggest the efficacy of pembrolizumab reported in the KEYNOTE-048 study is reflected in our real-world European population and is similar to other real-world studies in different geographical populations.

**Table 4.** Summary of efficacy outcomes from our study and the KEYNOTE-048 trial (total population group) [5]. DOR, duration of response; irAEs, immune-related adverse events; OS, overall survival; PFS, progression-free survival.

| Outcome | Scottish study |  | KEYNOTE-048 study |  |
| --- | --- | --- | --- | --- |
|  | Pembrolizumab monotherapy | Pembrolizumab -chemotherapy | Pembrolizumab monotherapy | Pembrolizumab -chemotherapy |
| OS - median (months) | 11.4 | Not yet reached | 11.6 | 13.0 |
| OS - 12 months | 45% | 60% | 49% | 53% |
| OS - 24 months | 27% | Not yet reached | 27% | 29% |
| PFS - 12 months | 22% | 20% | 17% | 17% |
| DOR - median (months) | 13.3 | 7.3 | 22.6 | 6.7 |

Patient characteristics in our study differed between those receiving pembrolizumab monotherapy and pembrolizumab-chemotherapy. A higher response rate and better early survival performance have been reported in the KEYNOTE-048 study for patients receiving the latter treatment [5]. Based on this, patients in daily clinical practice analysed in our study were selected for pembrolizumab-chemotherapy if there was rapidly growing and/or highly symptomatic disease and if they were able to tolerate the potential increased toxicity of the combination regimen. Due to this selection bias, we analysed patients who received pembrolizumab-chemotherapy as a separate population from those who received pembrolizumab monotherapy. Nonetheless, a higher rate of radiological response was seen in patients receiving pembrolizumab-chemotherapy compared with pembrolizumab monotherapy (Figure 2) primarily due to a lower proportion of mixed responses or stable disease. This qualified observation is in keeping with the findings of the KEYNOTE-048 study and supportive of the rationale behind the choice to offer combination treatment for select patient populations.

The benefit of being able to select pembrolizumab monotherapy or pembrolizumab-chemotherapy based on clinical factors has already been highlighted and appears to be essential in this patient group (as we described earlier) [11]. This, coupled with exploratory data suggesting that differing levels of PD-L1 expression may support different treatment strategies [12], means that access to both pembrolizumab monotherapy and in combination with chemotherapy is vital as we endeavour to optimally individualise treatment for our patients. At present, patients in England and Wales lack access to this option [10]. For those with a high disease burden or rapidly progressing disease, the only options remain either pembrolizumab monotherapy with lower ORR and potentially lower OS in the initial months, or cytotoxic chemotherapy with higher ORR that lacks the more durable responses seen with immunotherapy. The real-world data we present from Scotland therefore supports patients and colleagues in England and Wales in seeking NICE approval for pembrolizumab-chemotherapy.

Our cohort contained a substantial number (n=33) of patients with newly diagnosed, non-metastatic HNSCC (Table 1). This group made up a low proportion of the KEYNOTE-048 study population (n=7), thus we provide novel performance data in this subgroup [5]. Despite the substantially larger proportion of this subgroup in our cohort, our efficacy endpoints were comparable to the KEYNOTE-48 study. Indeed, our univariable analysis for patients receiving pembrolizumab monotherapy found no significant association between disease status and OS or PFS (Table 3). Our multivariable model found metastatic disease was significantly associated with worse PFS as compared with newly diagnosed, non-metastatic disease (Table 2), however, this analysis was not designed with the appropriate cofactors to specifically test this independent association. Overall, our analysis suggests pembrolizumab performs effectively in treatment-naïve patients with newly diagnosed, non-metastatic disease. Patients presenting with advanced locoregional disease are often difficult to manage with poor outcomes despite morbid, multi-modality radical treatment. Patients are sometimes unfit to undergo such intensive treatment or, knowing the morbidity and low success rates, patients sometimes opt for a palliative approach. Our study highlights that the principles of first-line palliative management for this patient subgroup are equivalent to those applied to the management of recurrent and metastatic disease.

A patient subgroup not represented in the KEYNOTE-048 study but present in our cohort were those where PS=2. The patients in our cohort had acutely deteriorated to PS2 due to recent disease factors, rather than chronic comorbidities. Thus, treatment with pembrolizumab was deemed to be appropriate despite poorer PS. Our univariable analysis found PS2 was associated with poorer OS, in keeping with previous studies investigating patients with HNSCC receiving immune checkpoint inhibitors [28, 29]. Given only five patients had a PS=2, further studies for this PS are needed and are currently underway [30].

We acknowledge that the toxicity profile reported in our cohort is focused on irAEs and somewhat differs from the KEYNOTE-048 study findings. Whilst grade ≥3 adverse events are likely to be recorded in routine clinical records, it remains less rigorous than the careful documentation of adverse events in clinical trials. This may explain the differences between our data and the KEYNOTE-048 study, thus direct toxicity comparisons must be interpreted with caution. However, like KEYNOTE-048, our results show that severe irAE toxicity is uncommon and this is a well-tolerated treatment in the real-world HNSCC population.

Experiencing irAEs when receiving PD-1/PD-L1 targeting immune checkpoint inhibitors has been associated with better clinical outcomes in multiple studies [16, 18, 23, 24, 31, 32]. We report that experiencing ≥1 irAE when receiving pembrolizumab monotherapy was associated with favourable OS and PFS versus experiencing no irAEs. This was maintained after controlling for possible confounding factors, by multivariable analysis, suggesting an independent association. We report a trend (not statistically significant) toward an association between irAEs and better survival outcomes when receiving pembrolizumab-chemotherapy. Given we found a strong association between higher DCR and experiencing ≥1 irAE in this group, a survival benefit would be expected. The small sample size within our cohort, particularly of evaluable patients at 24 months, may explain this lack of statistical significance. Thus, further follow-up of a larger cohort will help clarify associations within this group.

To our knowledge, the independent association between irAEs and better outcomes using multivariate analysis has not previously been reported for first-line pembrolizumab as a palliative treatment for HNSCC. Another real-world study, in a Japanese cohort, reported an association between experiencing any adverse event, not restricted to irAEs, with better OS for patients receiving pembrolizumab monotherapy in univariable analysis [26]. In contrast, analysis of real-world data in a different Japanese cohort found no association between irAEs and OS [25], though in a smaller cohort than our own.

Within our cohort, experiencing ≥1 irAE was associated with a markedly higher radiological response rate and DCR for patients receiving both pembrolizumab treatment options. This has been described previously in patients receiving nivolumab for HNSCC [16, 24]. We believe this is the first report of this association for first-line pembrolizumab as a palliative treatment for HNSCC.

A strength of our study was the analogous measurement of survival between our cohort and the KEYNOTE-048 study, both from the date of treatment initiation (or up to 5 days prior in KEYNOTE-048), thus providing comparable data. Despite the retrospective nature of our study, our outcomes are in keeping with those from the KEYNOTE-048 study and suggest our data is likely to be robust generally. A limitation of our study was that radiology did not explicitly use response criteria evaluation in solid tumours (RECIST) when reporting and so radiological response classification was determined by comparison of the radiology reported evaluations and the clinical evaluation of the treating clinician retrospectively [33]. We acknowledge that 21 patients (around 23%) of the whole cohort had no tumour PD-L1 status reflecting guidelines for the use of pembrolizumab during the COVID-19 pandemic. The KEYNOTE-048 included PDL1-positive and negative disease, with around 85% of tumours overexpressing PD-L1. We anticipate the same proportion in our cohort and have evaluated outcomes in our study against the whole KEYNOTE-048 population as this was felt to be the most equitable comparison. A further limitation of our study was the modest sample size. However, our numbers are similar to the larger of the previously published cohorts, but with European data and longer follow-up, therefore, offering new insights into the real-world performance of pembrolizumab in the longer term and data on its use in the newly-diagnosed, non-metastatic population.

## Conclusion

In conclusion, we performed a national, multi-centre, real-world, retrospective cohort study of patients receiving pembrolizumab with or without chemotherapy as a first-line palliative treatment for HNSCC. Our data shows pembrolizumab, with or without chemotherapy, performs comparably to the pivotal Phase 3 KEYNOTE-048 study, in day-to-day clinical practice, including in patients with newly diagnosed, non-metastatic disease. We also conclude that irAEs, as reported within routine clinical records, are highly associated with better clinical outcomes in patients receiving pembrolizumab monotherapy, with a trend towards an association in patients receiving pembrolizumab-chemotherapy.

Finally, despite real-world selection bias of rapid disease progression or high disease burden in the pembrolizumab-chemotherapy cohort, we report higher response rates and survival compared with pembrolizumab monotherapy, supporting routine access to both treatment options for clinicians and patients.

## Data Availability

All data produced in the present study are available upon reasonable request to the authors.

## Abbreviations

HNSCC: head and neck squamous cell carcinoma
irAE: immune-related adverse event
PD-L1: programmed death-ligand 1

## Acknowledgements

We thank Andrew Pulford (Public Health Scotland) for analysis approach discussion. Claire Paterson is supported by NHS Research Scotland (NRS) senior fellowship, Beatson Cancer Charity and CRUK RadNet Glasgow. David Noble is supported by an NHS Research Scotland (NRS) senior fellowship and the Jamie King Foundation.

## References

[1] Creaney G, McMahon AD, Ross AJ, Bhatti LA, Paterson C, Conway DI. Head and neck cancer in the UK: what was the stage before COVID-19? UK cancer registries analysis (2011-2018). Br Dent J. 2022;233:787–93.

[2] Machiels JP, Rene Leemans C, Golusinski W, Grau C, Licitra L, Gregoire V, et al. Squamous cell carcinoma of the oral cavity, larynx, oropharynx and hypopharynx: EHNS-ESMO-ESTRO Clinical Practice Guidelines for diagnosis, treatment and follow-up. Ann Oncol. 2020;31:1462–75.

[3] Public Health Scotland. Cancer survival statistics - People diagnosed with cancer during 2015 to 2019. 2022.

[4] Waldman AD, Fritz JM, Lenardo MJ. A guide to cancer immunotherapy: from T cell basic science to clinical practice. Nat Rev Immunol. 2020;20:651–68.

[5] Burtness B, Harrington KJ, Greil R, Soulieres D, Tahara M, de Castro G, Jr., et al. Pembrolizumab alone or with chemotherapy versus cetuximab with chemotherapy for recurrent or metastatic squamous cell carcinoma of the head and neck (KEYNOTE-048): a randomised, open-label, phase 3 study. Lancet. 2019;394:1915–28.

[6] National Comprehensive Cancer Network. Head and Neck Cancers Version 2.2022. 2022.

[7] Harrington KJ, Burtness B, Greil R, Soulieres D, Tahara M, de Castro G, Jr., et al. Pembrolizumab With or Without Chemotherapy in Recurrent or Metastatic Head and Neck Squamous Cell Carcinoma: Updated Results of the Phase III KEYNOTE-048 Study. J Clin Oncol. 2022:JCO2102508.

[8] Scottish Government. Coronavirus (COVID-19): interim governance framework for cancer medicines in adults. 2020.

[9] Scottish Medicines Consortium. Pembrolizumab 25mg/mL concentrate for solution for infusion and 50mg powder for concentrate for solution for infusion (Keytruda®). 2020.

[10] National Institute for Health and Care Excellence. Pembrolizumab for untreated metastatic or unresectable recurrent head and neck squamous cell carcinoma. 2020.

[11] Harrington KJ, Bhide SA, Forster MD, Good JS, Gunn L, Kong A, et al. Looking a Gift Horse in the Mouth: Observations on NHS England’s Interim Guidance on Pembrolizumab in Head and Neck Squamous Cell Cancer. Clin Oncol (R Coll Radiol). 2020;32:490–2.

[12] Burtness B, Rischin D, Greil R, Soulieres D, Tahara M, de Castro G, Jr., et al. Pembrolizumab Alone or With Chemotherapy for Recurrent/Metastatic Head and Neck Squamous Cell Carcinoma in KEYNOTE-048: Subgroup Analysis by Programmed Death Ligand-1 Combined Positive Score. J Clin Oncol. 2022;40:2321–32.

[13] Pasello G, Pavan A, Attili I, Bortolami A, Bonanno L, Menis J, et al. Real world data in the era of Immune Checkpoint Inhibitors (ICIs): Increasing evidence and future applications in lung cancer. Cancer Treat Rev. 2020;87:102031.

[14] Cramer-van der Welle CM, Verschueren MV, Tonn M, Peters BJM, Schramel F, Klungel OH, et al. Real-world outcomes versus clinical trial results of immunotherapy in stage IV non-small cell lung cancer (NSCLC) in the Netherlands. Sci Rep. 2021;11:6306.

[15] Khozin S, Miksad RA, Adami J, Boyd M, Brown NR, Gossai A, et al. Real-world progression, treatment, and survival outcomes during rapid adoption of immunotherapy for advanced non-small cell lung cancer. Cancer. 2019;125:4019–32.

[16] Matsuo M, Yasumatsu R, Masuda M, Toh S, Wakasaki T, Hashimoto K, et al. Relationship between immune-related adverse events and the long-term outcomes in recurrent/metastatic head and neck squamous cell carcinoma treated with nivolumab. Oral Oncol. 2020;101:104525.

[17] Haratani K, Hayashi H, Chiba Y, Kudo K, Yonesaka K, Kato R, et al. Association of Immune-Related Adverse Events With Nivolumab Efficacy in Non-Small-Cell Lung Cancer. JAMA Oncol. 2018;4:374–8.

[18] Freeman-Keller M, Kim Y, Cronin H, Richards A, Gibney G, Weber JS. Nivolumab in Resected and Unresectable Metastatic Melanoma: Characteristics of Immune-Related Adverse Events and Association with Outcomes. Clin Cancer Res. 2016;22:886–94.

[19] Cortellini A, Friedlaender A, Banna GL, Porzio G, Bersanelli M, Cappuzzo F, et al. Immune-related Adverse Events of Pembrolizumab in a Large Real-world Cohort of Patients With NSCLC With a PD-L1 Expression >/= 50% and Their Relationship With Clinical Outcomes. Clin Lung Cancer. 2020;21:498–508 e2.

[20] Benchimol EI, Smeeth L, Guttmann A, Harron K, Moher D, Petersen I, et al. The REporting of studies Conducted using Observational Routinely-collected health Data (RECORD) statement. PLoS Med. 2015;12:e1001885.

[21] Chennamadhavuni A, Abushahin L, Jin N, Presley CJ, Manne A. Risk Factors and Biomarkers for Immune-Related Adverse Events: A Practical Guide to Identifying High-Risk Patients and Rechallenging Immune Checkpoint Inhibitors. Front Immunol. 2022;13:779691.

[22] Okada T, Fushimi C, Matsuki T, Tokashiki K, Takahashi H, Okamoto I, et al. Effects of Pembrolizumab in Recurrent/Metastatic Squamous Cell Head and Neck Carcinoma: A Multicenter Retrospective Study. Anticancer Res. 2023;43:2717–24.

[23] Ishihara H, Takagi T, Kondo T, Homma C, Tachibana H, Fukuda H, et al. Association between immune-related adverse events and prognosis in patients with metastatic renal cell carcinoma treated with nivolumab. Urol Oncol. 2019;37:355 e21–e29.

[24] Okamoto I, Sato H, Kondo T, Koyama N, Fushimi C, Okada T, et al. Efficacy and safety of nivolumab in 100 patients with recurrent or metastatic head and neck cancer - a retrospective multicentre study. Acta Otolaryngol. 2019;139:918–25.

[25] Sano D, Tokuhisa M, Takahashi H, Hatano T, Nishimura G, Ichikawa Y, et al. Real-world Therapeutic Outcomes of the Pembrolizumab Regimen as First-line Therapy for Recurrent/Metastatic Squamous Cell Carcinoma of the Head and Neck: A Single-center Retrospective Cohort Study in Japan. Anticancer Res. 2022;42:4477–84.

[26] Nakano T, Yasumatsu R, Hashimoto K, Kuga R, Hongo T, Yamamoto H, et al. Real-world Experience With Pembrolizumab for Advanced-stage Head and Neck Cancer Patients: A Retrospective, Multicenter Study. Anticancer Res. 2022;42:3653–64.

[27] Yalamanchali A, Yang K, Roof L, Lopetegui-Lia N, Schwartzman LM, Campbell SR, et al. Comparison of real-world outcomes following immunotherapy in recurrent or metastatic head and neck squamous cell carcinoma with outcomes of randomized controlled trials. Head Neck. 2023;45:862–71.

[28] Chalker C, Voutsinas JM, Wu QV, Santana-Davila R, Hwang V, Baik CS, et al. Performance status (PS) as a predictor of poor response to immune checkpoint inhibitors (ICI) in recurrent/metastatic head and neck cancer (RMHNSCC) patients. Cancer Med. 2022;11:4104–11.

[29] Minohara K, Matoba T, Kawakita D, Takano G, Oguri K, Murashima A, et al. Novel Prognostic Score for recurrent or metastatic head and neck cancer patients treated with Nivolumab. Sci Rep. 2021;11:16992.

[30] Forsyth S, Yip K, Foran B, Gougis P, Wheeler G, White L, et al. 979TiP POPPY: A phase II trial to assess the efficacy and safety profile of pembrolizumab in patients with performance status 2 with recurrent or metastatic squamous cell carcinoma of the head and neck. Annals of Oncology. 2020;31:S686–S7.

[31] Michot JM, Bigenwald C, Champiat S, Collins M, Carbonnel F, Postel-Vinay S, et al. Immune-related adverse events with immune checkpoint blockade: a comprehensive review. Eur J Cancer. 2016;54:139–48.

[32] Ricciuti B, Genova C, De Giglio A, Bassanelli M, Dal Bello MG, Metro G, et al. Impact of immune-related adverse events on survival in patients with advanced non-small cell lung cancer treated with nivolumab: long-term outcomes from a multi-institutional analysis. J Cancer Res Clin Oncol. 2019;145:479–85.

[33] Park HJ, Kim GH, Kim KW, Lee CW, Yoon S, Chae YK, et al. Comparison of RECIST 1.1 and iRECIST in Patients Treated with Immune Checkpoint Inhibitors: A Systematic Review and Meta-Analysis. Cancers (Basel). 2021;13.

