## Supplementary Table S1 and Table S2 for "The UK Divide: Does having a Pembrolizumab-Chemotherapy option in head and neck cancer matter? Real-world experience of first-line palliative pembrolizumab monotherapy and pembrolizumab-chemotherapy combination in Scotland"

Table S1. Any-grade immune-related adverse event profile. GI, gastrointestinal; irAE, immune-related adverse event.

| | Pembrolizumab monotherapy | | Pembrolizumab -chemotherapy | |
|---|---|---|---|---|
| | n = 76 | | n = 15 | |
| irAE, n (%) | Any grade | Grade ≥3 | Any grade | Grade ≥3 |
| GI disorders | 16 (21.1) | 4 (5.3) | 1 (6.7) | 1 (6.7) |
| Hepatitis | 6 (7.9) | 3 (3.9) | 0 (0.0) | 0 (0.0) |
| Diarrhoea | 8 (10.5) | 1 (1.3) | 1 (6.7) | 1 (6.7) |
| Oesophagitis | 1 (1.3) | 0 (0.0) | 0 (0.0) | 0 (0.0) |
| Mucositis | 1 (1.3) | 0 (0.0) | 0 (0.0) | 0 (0.0) |
| Endocrine disorders | 14 (18.4) | 2 (2.6) | 1 (6.7) | 0 (0.0) |
| Hypothyroidism | 11 (14.5) | 0 (0.0) | 1 (6.7) | 0 (0.0) |
| Hypoadrenalism | 2 (2.6) | 2 (2.6) | 0 (0.0) | 0 (0.0) |
| Hyperthyroidism | 1 (1.3) | 0 (0.0) | 0 (0.0) | 0 (0.0) |
| Systemic, n (%) | 9 (11.8) | 0 (0.0) | 3 (20.0) | 0 (0.0) |
| Fatigue | 9 (11.8) | 0 (0.0) | 3 (20.0) | 0 (0.0) |
| Dermatological disorders | 5 (6.6) | 1 (1.3) | 2 (13.3) | 1 (6.7) |
| Rash | 3 (3.9) | 1 (1.3) | 2 (13.3) | 1 (6.7) |
| Pruritus | 1 (1.3) | 0 (0.0) | 0 (0.0) | 0 (0.0) |
| Peri-oral rash | 1 (1.3) | 0 (0.0) | 0 (0.0) | 0 (0.0) |
| Musculoskeletal disorders | 4 (5.3) | 0 (0.0) | 1 (6.7) | 0 (0.0) |
| Arthralgia | 4 (5.3) | 0 (0.0) | 1 (6.7) | 0 (0.0) |
| Respiratory disorders | 1 (1.3) | 0 (0.0) | 1 (6.7) | 0 (0.0) |
| Pneumonitis | 1 (1.3) | 0 (0.0) | 1 (6.7) | 0 (0.0) |
| Renal disorders | 0 (0.0) | 0 (0.0) | 1 (6.7) | 0 (0.0) |
| Nephritis | 0 (0.0) | 0 (0.0) | 1 (6.7) | 0 (0.0) |
| Neurological disorders | 1 (1.3) | 0 (0.0) | 0 (0.0) | 0 (0.0) |
| Neuropathic pain | 1 (1.3) | 0 (0.0) | 0 (0.0) | 0 (0.0) |

Table S2. Palliative radiotherapy received before or during pembrolizumab therapy. Each line represents an individual patient. LNs, lymph nodes; mets, metastases; NR, data not retrieved from or not recorded in clinical records; P, pembrolizumab-containing therapy.

| Pembrolizumab treatment received | Palliative radiotherapy 1 | | | Palliative radiotherapy 2 | | | Palliative radiotherapy 3 | | | Palliative radiotherapy 4 | | |
|---|---|---|---|---|---|---|---|---|---|---|---|---|
| | Time | Dose* | Site | Time | Dose* | Site | Time | Dose* | Site | Time | Dose* | Site |
| Monotherapy | During P | 10/1 | Regional LNs | During P | 8/1 | Regional LNs | During P | 20/5 | Regional LNs | | | |
| Monotherapy | During P | 30/10 | Distant mets | | | | | | | | | |
| Monotherapy | Before P | 8/1 | Primary & Regional LNs | | | | | | | | | |
| Monotherapy | During P | 20/5 | Primary & Regional LNs | | | | | | | | | |
| Monotherapy | During P | 8/1 | Regional LNs | | | | | | | | | |
| Monotherapy | Before P | 50/25 | Regional LNs | | | | | | | | | |
| Monotherapy | Before p | 8/1 | Distant mets | Before P | 8/1 | Distant mets | During P | 8/1 | Distant mets | During P | 8/1 | Distant mets |
| Monotherapy | Before P | NR | NR | | | | | | | | | |
| Monotherapy | Before P | 20/5 | NR | | | | | | | | | |
| Monotherapy | Before P | 8/1 | NR | | | | | | | | | |
| Monotherapy | Before P | 20/5 | Primary & Regional LNs | During P | 8/1 | Primary & Regional LNs | | | | | | |
| Monotherapy | Before P | 8/1 | Distant mets | During P | 8/1 | Regional LNs & Distant mets | | | | | | |
| Monotherapy | Before P | 65/30 | Primary & Regional LNs | Before P | 54/3 | Lung | | | | | | |
| Monotherapy | Before P | 30/10 | Primary | | | | | | | | | |
| Monotherapy | Before P | 30/10 | Primary | | | | | | | | | |
| Monotherapy | Before P | 30/15 | Primary | | | | | | | | | |
| Monotherapy | Before P | 30/10 | Primary & Regional LNs | Before P | 20/10 | Primary & Regional LNs | | | | | | |

| | | | |
|---|---|---|---|
| Monotherapy | Before IO | 30/15 | Primary |
| Monotherapy | During IO | 30/10 | Distant Mets |
| With chemotherapy | During IO | 65/30 | Primary |

**Dose in gy/number of fractions*
